# Multi-omic and Survival Analysis of Acute Chest Syndrome Burden in Sickle Cell Disease

**DOI:** 10.64898/2026.10.15.26365641

**Authors:** Anvita Kulshrestha, Melanie E. Garrett, Seyed Mehdi Nouraie, Roberto F. Machado, Victor R. Gordeuk, Mark T. Gladwin, Marilyn J. Telen, Allison E. Ashley-Koch, NHLBI Trans-Omics for Precision Medicine (TOPMed)

## Abstract

Sickle cell disease (SCD) results from homozygosity or compound heterozygosity for a single point mutation in the beta hemoglobin gene. Despite carrying the same underlying causal mutation, patients exhibit highly heterogeneous clinical outcomes. Acute chest syndrome (ACS) is one of the leading causes of mortality in SCD. In this study, we aimed to identify genetic variants associated with ACS occurrence and recurrence (>5 episodes) in two SCD cohorts (OMG-SCD and Walk-PHaSST) of predominantly African ancestry and characterize their downstream molecular consequences. Although none of the SNPs reached the genome-wide threshold in the occurrence meta-analysis, we identified 40 independent genetic loci with p <10^-5^. However, the recurrence GWAS in OMG-SCD, which compared individuals with no prior history of ACS to individuals with a history of >5 episodes, identified one genome-wide significant locus along with 38 other independent loci with p <10^-5^. Survival analysis suggested lower survival probability among young ACS recurrence cases compared to controls or occurrence cases. Furthermore, molecular QTL analysis of ACS recurrence identified 42 SNP-protein and eight SNP-metabolite associations with p <10^-3^. Pathway and enrichment analysis identified secretory, platelet/hemostatic, and complement-mediated pathways, confirming the complex and heterogeneous pathophysiology of ACS. This study highlights the importance of a well-defined phenotype for identification of risk loci and elucidates the complex interplay of hemolytic, inflammatory, and cellular organization pathways that warrants further evaluation through large-scale multi-omic studies of SCD patients. Ultimately, the more extreme phenotype of ACS recurrence may be a more informative phenotype for analysis, risk prediction, and therapeutic targeting.

## INTRODUCTION

Sickle cell disease (SCD) results from a homozygous or compound heterozygous point mutation in the beta hemoglobin (Hb) gene where glutamic acid at position 6 of the beta-globin chain is substituted with valine, resulting in the sickling of deoxygenated red blood cells (HbS genotype).^1^ Despite the same underlying causal mutation, patients exhibit highly heterogeneous clinical outcomes.^2^ Acute chest syndrome (ACS) is the leading cause of mortality among individuals with SCD, accounting for 25% of all deaths, and is the second most common reason for hospitalizations after vaso-occlusive pain.^3,4^ ACS is associated with extended hospital stays, risk of respiratory failure, and the potential for developing chronic lung disease after multiple episodes.^5–7^ The incidence of ACS is higher in children (< 5 years of age) than adults, as children are more susceptible to infections.^7,8^ However, adult cases of ACS tend to be more severe, associated with pain, and have higher rates of mortality, with a higher incidence of pulmonary fat and bone marrow emboli.^8–11^

The pathophysiology of ACS is complex and can develop for multiple reasons^12^: Bone marrow fat embolization may occur in up to one quarter of ACS episodes and is often found in the presence of concomitant infection;^9^ in most instances, the presence of fat emboli is suggested by clinical and hematologic characteristics rather than the diagnostic feature of fat laden pulmonary macrophages, whose detection requires bronchoscopy and bronchoalveolar lavage.^7,8^ At least half of ACS episodes occur without an identifiable cause but are often preceded by an acute vaso-occlusive crisis, suggesting that intravascular pulmonary sequestration of red blood cells (RBCs), neutrophils and platelets can lead to lung injury and infarction.^13^ Histology often reveals diffused alveolar damage, and elevated markers of inflammation both in the circulation and within bronchoalveolar lavage fluid are seen in patients with ACS.^14^ Primary pulmonary infection leading to ACS is most often identifiable in children.^8^ Influenza and COVID-19 are among the common viruses that can trigger ACS.^9,15^ Finally, a history of asthma, especially in children and adolescents, has been identified as a risk factor.^8^ Childhood asthma is associated with more frequent vaso-occlusive events, earlier onset of ACS, recurrent ACS, and mortality.^3,16,17^ In the absence of identifiable infection, the mainstays of treatment for ACS are RBC transfusion and empiric antibiotics for community-acquired pneumonia, as well as analgesics and oxygen supplementation when required.

There have been numerous attempts to identify genetic loci associated with ACS, mostly through candidate gene-based studies. Using the Comprehensive Study of Sickle Cell Disease (CSSCD) cohort, an intergenic SNP (rs6141803) between *COMMD7* and *DNMT3B* on chromosome 20 was associated with ACS but only trended in the right direction upon replication.^8,18^ COMMD7 is highly expressed in the lung and interacts with NF-κB signaling.^18^ Other studies have found genetic polymorphisms in genes within the nitric oxide (NO) metabolism and TGF-β pathways as risk factors for ACS.^19–23^ NO is thought to play a central role in vaso-occlusive crisis pathogenesis,^24^ as hemolysis results in the release of cell-free hemoglobin, which rapidly consumes NO.^25^ Reduced NO bioavailability may then lead to endothelial injury, leukocyte adhesion and downstream ischemia.^24,25^ The TGF-β pathway is known to play a role in immune response, angiogenesis, inflammation, hematopoiesis, vascular inflammation, and cell proliferation.^26^ In a candidate gene-based study, Bean et al. identified polymorphisms in the heme oxygenase 1 (*HMOX1*) promoter region that resulted in decreased rates of hospitalization for ACS.^27^*HMOX1* produces the protein HO-1, which is the rate-limiting enzyme in the catabolism of heme, and is thought to attenuate the severity of clinical outcomes as a result of hemolytic crisis.^27,28^ *HMOX1* induction has been shown to be cytoprotective against hemolysis and vaso-occlusive crisis in murine SCD models.^29^ However, genome-wide association studies (GWAS) have failed to replicate the results of targeted genetic associations or to conclusively identify risk loci for ACS, likely due to its multifactorial and complex etiology.^8^ Moreover, the genetic architecture of ACS recurrence, which we hypothesize carries clinical risk distinct from that of a first event, remains largely uncharacterized.

To address this gap, we conducted a meta-analysis across the two of the largest available adult SCD cohorts in the US - Outcome Modifying Genes in SCD (OMG-SCD) and Walk-Treatment of Pulmonary Hypertension and SCD with Sildenafil Therapy (Walk-PHaSST) - to identify variants associated with ACS occurrence, followed by a GWAS for ACS recurrence (>5 episodes) in the OMG-SCD cohort. We then tested top recurrence-associated SNPs for independent association with plasma protein and metabolite abundance in OMG-SCD and mapped the nearest genes onto protein-protein interaction networks to identify shared biological pathways connecting genetic variation to the molecular phenotypes.

## MATERIALS AND METHODS

### Participants and Endpoints

Adult (age ≥18 years) SCD participants with genetic data and medical history inclusive of ACS events were selected from two previously described cohorts for inclusion in this study. <u>Primary dataset:</u> 430 participants with severe SCD genotype (HbSS or HbSβ^0^) and complete covariate data (age, sex, hydroxyurea status) were included from OMG-SCD^30^, a multi-center cohort from the southeastern US, ascertained for a cross-sectional observational study of genetic modifiers of SCD. In this cohort, ACS was defined as a constellation of signs and symptoms, including fever, leukocytosis, pulmonary infiltrate(s), chest pain, and hypoxemia. A smaller subset of participants (n=192) was selected based on extreme phenotype (> 5 lifetime episodes of ACS vs none) for the recurrence GWAS. Patients with >5 episodes of ACS during lifetime were classified as cases (n=80), while patients with no history of ACS were classified as controls (n=112).

<u>Second dataset</u>: 275 participants were included from the Walk-PHaSST study, a clinical trial of sildenafil, for which participants were enrolled during the screening phase at centers in the US and United Kingdom.^31^ The definition of ACS in this cohort was similar to that in OMG-SCD. In both cohorts, diagnosis of pneumonia was considered ACS.

All participants gave informed consent and study protocols were approved by local institutional review boards or ethics committees. Participants provided blood samples for genetic analyses at study enrollment, and a variety of clinical variables were obtained at steady state.

### Whole genome sequencing

Whole genome sequencing (∼30X) was performed on DNA isolated from whole blood at Baylor College of Medicine under the NHLBI TOPMed program. All samples included in this study are from TOPMed freeze 8, and data processing and primary quality control (QC) were performed at the Informatics Research Center (IRC) at the University of Michigan, methods for which have been previously described.^32,33^ In summary, reads were mapped to the GRCh38 assembly, variants were jointly called across roughly 158,000 freeze 8 samples, and a support vector machine classifier was applied to filter the resulting call set. Samples were further screened for discordant sex assignment, inconsistencies with reported pedigree, and agreement with prior array-based genotypes. We retained only variants covered at ≥10X read depth. OMG-SCD and Walk-PHaSST were each subjected to a second, cohort-specific round of QC in which we dropped variants with call rates below 85% or minor allele frequency (MAF) <1% and kept only biallelic single nucleotide polymorphisms (SNPs). Relatedness was then assessed by computing identity-by-descent (IBD) estimates in PLINK on a linkage disequilibrium (LD)-pruned set of SNPs with MAF >20%. For any pair with an IBD estimate above 0.25 (corresponding to second-degree relatedness), one member was dropped at random; this removed 31 Walk-PHaSST samples (n=275), while OMG-SCD contained no pairs related at the second degree or closer (n=430).

### Statistical analysis

Differences in participant characteristics between OMG-SCD and Walk-PHaSST as well as within OMG-SCD participants were analyzed in R using logistic regression (*glm, family = binomial*) for categorical variables and linear regression (*glm, family = gaussian*) for continuous variables. Survival curves were modeled and plotted using *survival* and *survminer* R packages, respectively.^34,35^ Covariate-adjusted survival was modeled using a Cox proportional hazards test. GWAS was performed using the R package SAIGE for both the ACS occurrence and recurrence analyses.^36^ Age, sex, hydroxyurea (HU) status and genomic PCs were included as covariates in both analyses. Two principal components (PCs) in OMG-SCD and five PCs in Walk-PHaSST were sufficient to account for population substructure. Fixed effects meta-analysis was accomplished with METAL^37^, combining ACS occurrence GWAS results across the two cohorts comprising 492 cases/213 controls (N=705) and 9,996,265 variants that passed quality control and were present in both cohorts. For the recurrence GWAS, association testing was performed in 80 cases and 112 controls across 8,639,124 variants. Using standard thresholds, SNPs with p-value<5x10^-8^ were considered genome-wide significant and those with p<10^-5^ were considered nominally significant. Quantile-quantile (QQ) and Manhattan plots were produced using *qqman* in R.^38^ LD clumping was done using PLINK 1.9 to identify independent lead SNPs.^39^ Genetic variants were annotated using ANNOVAR with hg38 reference built using dbSNP147 identifiers.^40^

Molecular quantitative trait loci (QTL) analysis was performed in R using generalized linear models (*glm*). Nontargeted plasma proteomic data^41^ as well as metabolomic data^42^ were generated as described in our earlier publications. False discovery rate (FDR) q-values for molecular QTL analysis were generated using *qvalue* in R.^43^ Protein-protein interaction network was generated using the STRING database (v12.0, Homo sapiens, medium confidence threshold - > 0.4).^44^ The network consisted of genes/nearest genes corresponding to all the proteins and SNPs that were nominally significant (p<0.001). This resulted in 54 candidate genes (excluding pseudogenes, non-coding RNA, and genes not in the STRING database). Genes with no interaction partners at this threshold were excluded from the visualization to focus on the connected subnetwork, leading to 30 genes being visualized. To characterize the biological roles represented within the connected network, functional enrichment was performed using STRING’s built-in enrichment tool.

## RESULTS

Participant characteristics by cohort are shown in Table 1. OMG-SCD participants were younger (33.55 vs. 36.96 years old, p=2.55x10^-4^), had a lower proportion of patients on HU (41.16% vs. 50.54%, p=0.015), and had a higher proportion of ACS occurrence (73.95% vs. 63.27%, p=0.003) than Walk-PHaSST participants included in the meta-analysis.

**Table 1:** Comparison of participant characteristics in the two cohorts used for ACS occurrence meta-analysis and among ACS groups within OMG-SCD patients.

|  | ACS Occurrence Meta-analysis |  |  | ACS Recurrence GWAS in OMG-SCD |  |  |  |  |  |
| --- | --- | --- | --- | --- | --- | --- | --- | --- | --- |
| Variable | OMG-SCD<br>(N = 430) | Walk-PHaSST<br>(N = 275) | p-value | Controls<br>(0 episodes) | Cases<sup>#</sup> (1-5 episodes) | Extreme Cases (>5 episodes) | p-value<br>(controls vs cases) | p-value<br>(controls vs extreme cases) | p-value<br>(cases vs extreme cases) |
| Female | 240<br>(55.81%) | 147<br>(53.45%) | 0.539 | 59<br>(52.68%,<br>N=112) | 135<br>(60.54%,<br>N=223) | 38 (47.50%,<br>N=80) | 0.170 | 0.479 | 0.044* |
| ACS Cases | 318<br>(73.95%) | 174<br>(63.27%) | 0.003* | - | - | - | - | - | - |
| On Hydroxyurea | 177<br>(41.16%) | 139<br>(50.54%) | 0.015* | 43<br>(38.39%,<br>N=112) | 85<br>(38.12%,<br>N=223) | 41 (51.25%,<br>N=80) | 0.961 | 0.078 | 0.042* |
| Age, years (Mean, SD) | 33.55<br>(12.12) | 36.96<br>(11.86) | 2.55 x<br>10<sup>-4</sup>* | 32.67<br>(12.28) | 34.35<br>(12.51) | 32.74<br>(11.00) | 0.233 | 0.970 | 0.309 |
| **Vaso-occlusive crisis requiring hospitalization** | - | - | - | 51<br>(46.79%,<br>N=109) | 130<br>(61.61%,<br>N=211) | 57 (75.00%,<br>N=76) | 0.011* | 1.76 x 10<sup>-4</sup>* | 0.037* |
| **Mortality** | - | - | - | 32<br>(51.61%,<br>N=62) | 54<br>(42.52%,<br>N=127) | 29 (54.71%,<br>N=53) | 0.239 | 0.740 | 0.136 |

SD: standard deviation, \* denotes p-value < 0.05, # Cases were excluded in recurrence GWAS but, were combined with extreme cases in occurrence meta-analysis.

### Meta-analysis identifies SNPs associated with occurrence of ACS

Meta-analysis identified 72 loci nominally associated with occurrence of ACS in OMG-SCD and Walk-PHaSST cohorts (λ = 1.03, Figure S1). After LD clumping, there were 40 independent loci nominally associated with occurrence of ACS (Table 2). Although none of the associations were genome-wide significant, our top hit was an intergenic SNP (rs143617547, p=2.34 x 10^-7^, Figure 1) on chromosome 2, between *GACAT3* and *FAM49A* genes. *GACAT3* lncRNA plays a tumor promoting role in lung cancer by activating the NF-kB and STAT3 signaling pathways.^45^ NF-kB has been thought to play a role in the inflammatory cascade during vaso-occlusive crisis, while STAT3 silencing has been linked to fetal hemoglobin activation as a means of inhibiting sickle polymerization.^46,47^ Among the proteins encoded by the closest genes mapped to the top five SNPs, NTRK3 (rs77723962) plays an important role in neurotrophin signaling in pulmonary inflammation, and UBR2 (rs16895923) triggers the activation of the NLRP1b inflammasome that drives inflammatory response leading to acute lung injury, supporting pulmonary inflammation as one of the major contributors to ACS.^48,49^

**Figure 1:**
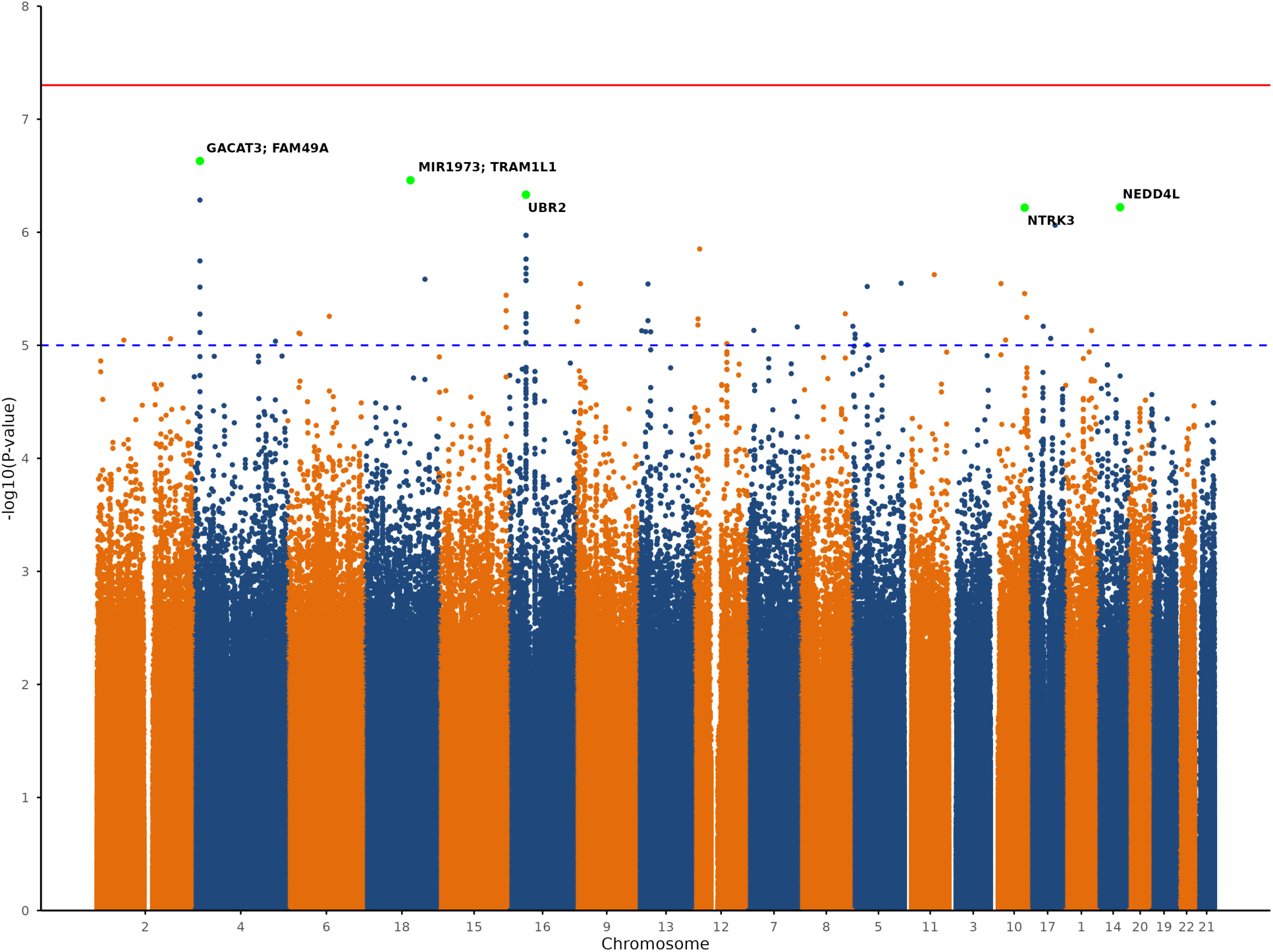
Manhattan plot of meta-analysis results for ACS occurrence in two SCD cohorts. Each point represents a SNP, ordered by chromosomal position (x-axis) and plotted against the −log₁₀ of its meta-analysis p-value (y-axis). The red solid line denotes genome-wide significance threshold (p = 5 x 10-8) and the blue dashed line is the suggestive threshold (p = 1 x 10-5). Top 5 independent SNPs have been labelled with their nearest genes.

**Table 2:**
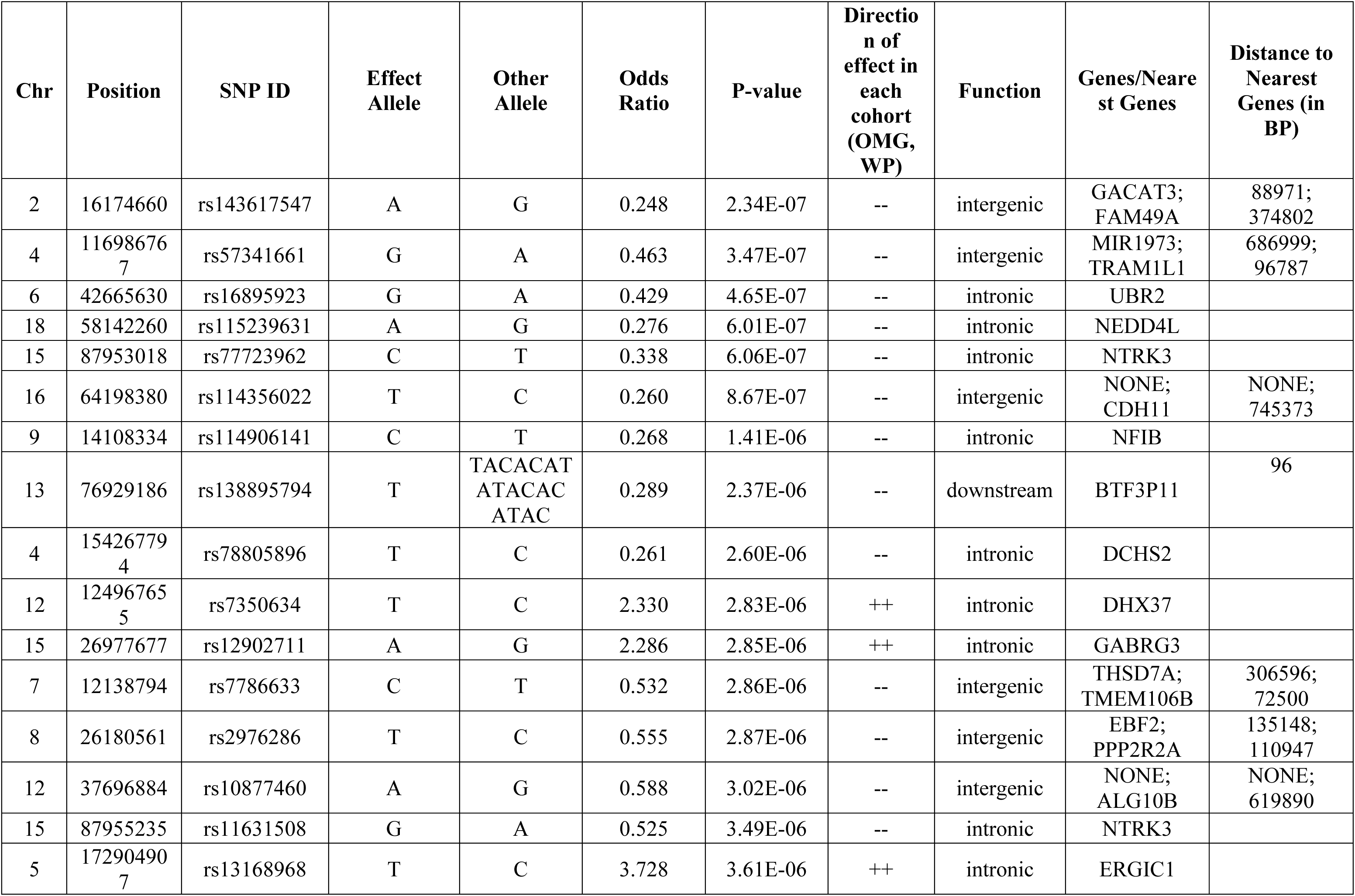
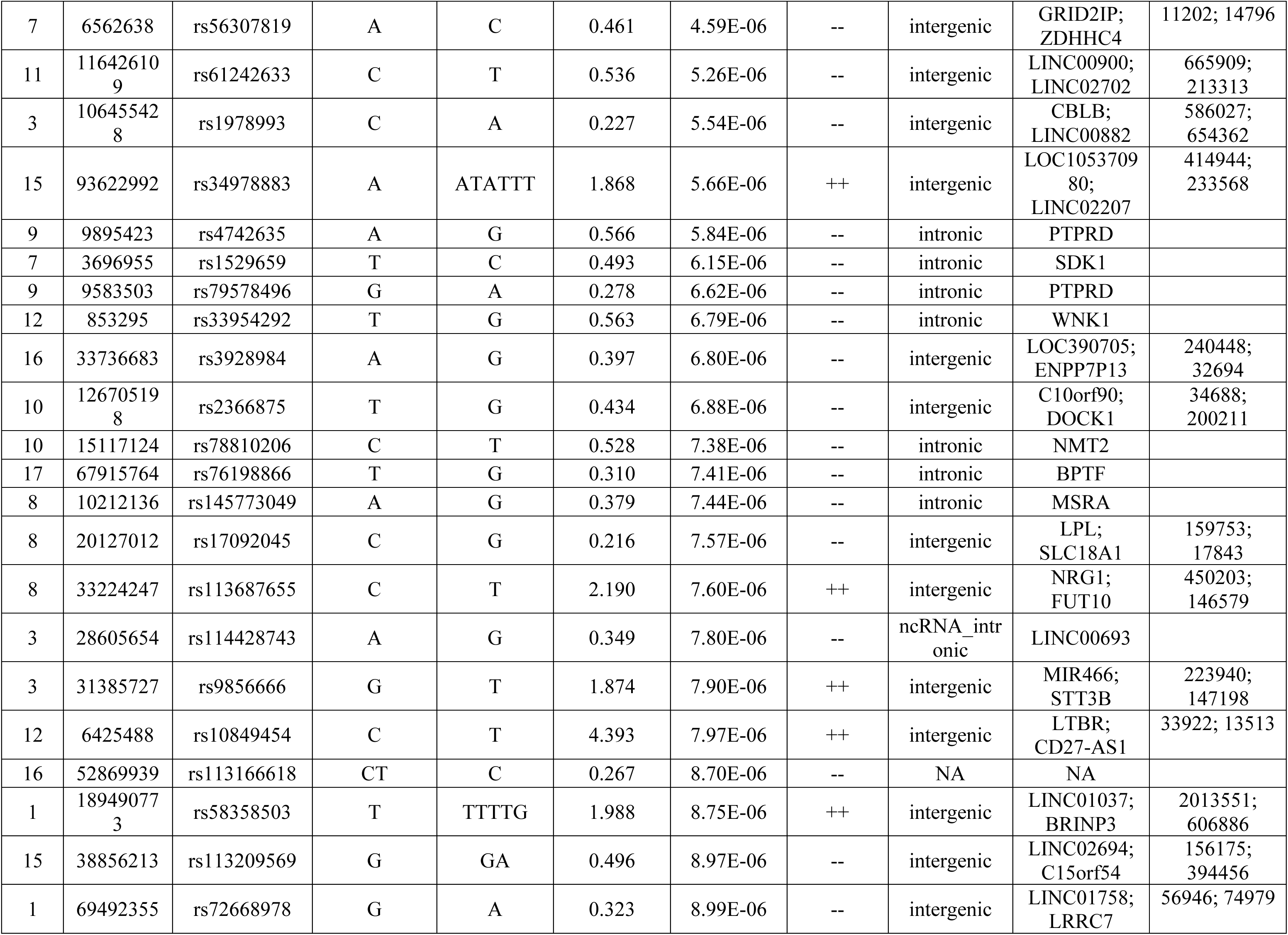
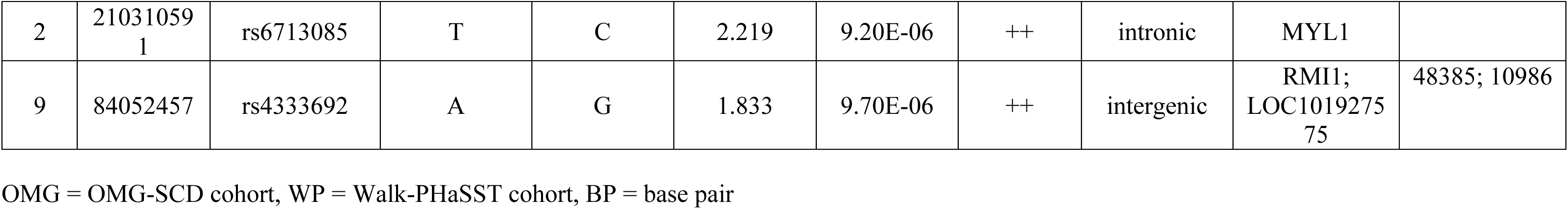
Top SNPs from ACS occurrence meta-analysis (p < 1 x 10^-5^).

### Distinct characteristics of extreme ACS cases

Because the meta-analysis did not identify genome-wide significant associations, we sought to reduce heterogeneity in the phenotype by focusing on the most severe cases with recurrent ACS. However, this analysis was limited to the OMG-SCD cohort, as the number of ACS events was only available in that cohort. Comparison of characteristics among controls (zero ACS episodes in lifetime), cases (1-5 ACS episodes in lifetime), and extreme cases (>5 ACS episodes in lifetime) is shown in Table 1. The extreme case group had a significantly lower percentage of females (47.50% vs 60.54%, p=0.044) and higher percentage of patients on hydroxyurea (51.25% vs 38.12%, p=0.042) as compared to the cases. Frequency of vaso-occlusive crisis requiring hospitalization was increased in patients with more ACS episodes, with extreme cases having the highest percentage of hospitalizations in the past year (75.00%, p(compared to controls)= 1.76 x 10^-4^, p(compared to cases)=0.037). Mortality was not significantly associated with ACS in our dataset (p=0.29, Figure 2A), even after adjusting for patients’ age at enrollment, sex, and hydroxyurea status. Interestingly, when we plotted survival curves stratified by age quantiles, we found that younger (aged 18-23 years) extreme cases had a much lower probability of survival as compared to controls and cases within the same age bracket across the ∼25 years follow-up (Figure 2B, Figure S2).

**Figure 2:**
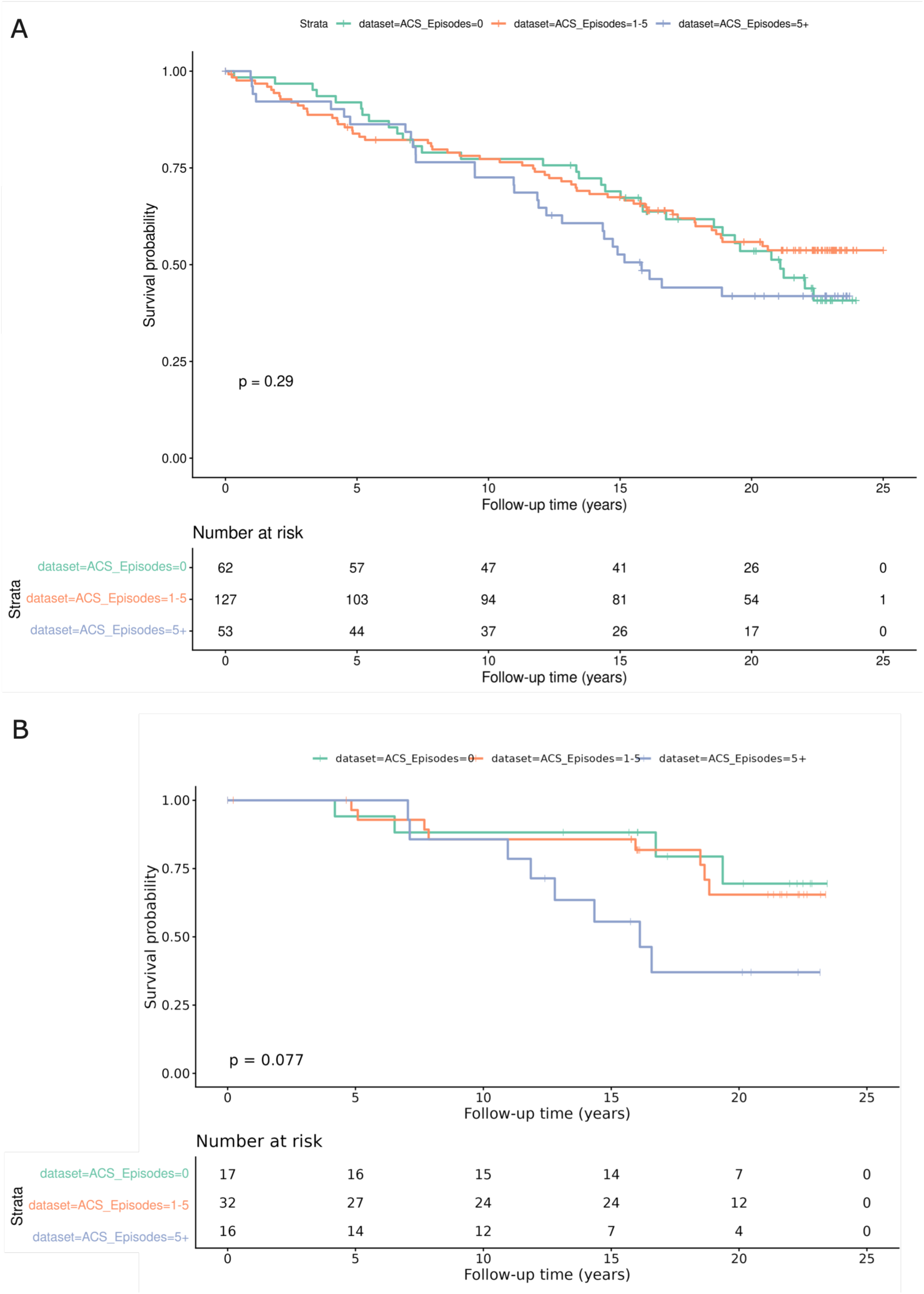
Survival curves for OMG-SCD participants. **(A)** Unadjusted survival curves for participants with 0 (n=62), 1-5 (n=127), and > 5 (n=53) ACS episodes. Vertical tick marks denote censored observations; the table below the plot gives the number at risk in each stratum at 5-year intervals. Groups did not differ significantly (log rank p=0.29). **(B):** Survival curve for the same three strata within the first quantile of age at examination: 18.0-23.0 years (0 episodes, n=17; 1-5, n=32; >5, n=16). Vertical tick marks denote censored observations and the table beneath gives the number remaining at risk at 5-year intervals. Log-rank p-values are also shown within each plot and were not adjusted for multiple comparisons.

### GWAS identifies a novel locus at chromosome 13 associated with recurrent ACS

The GWAS results for recurrent ACS (n = 192) are shown in Figure 3 (λ = 1.04, Figure S3). We identified 39 independent loci associated with ACS recurrence at a p-value < 1x10^-5^ (Table 3). Of these, 25 SNPs were also nominally associated in the ACS occurrence meta-analysis (p<0.05), 17 of which displayed the same direction of effect across both cohorts (Table S1). The top ACS recurrence-associated SNP was rs7319269 on chromosome 13 (Figure 3, Table 3), which reached genome-wide significance (p = 3.89x10^-8^). This intergenic SNP lies between *SNORA107* and *LINC00375,* both of which give rise to noncoding RNAs, which as a class have a broad role in protein expression and cellular homeostasis.^50^ Interestingly, another one of the top SNPs (rs1356744, an intronic SNP in *LINC01876* on chromosome 2, p = 1.02 x 10^-6^) has been previously associated with severe acute respiratory syndrome in COVID-19, hinting at a common biological pathway between ACS and COVID-19.^51^ Multiple SNPs in the same gene have also been shown to be associated with lung function.^52–58^ A recent study also showed that overexpression of LINC01876 is a potential biomarker for lung cancer, as it activates the AKT and ERK1/2 pathways.^59^ ERK 1/2 activation has been shown to trigger phosphorylation of cytoskeletal proteins and the adhesion molecule ICAM-4, promoting sickle RBC adhesion to the endothelium.^60^

**Figure 3:**
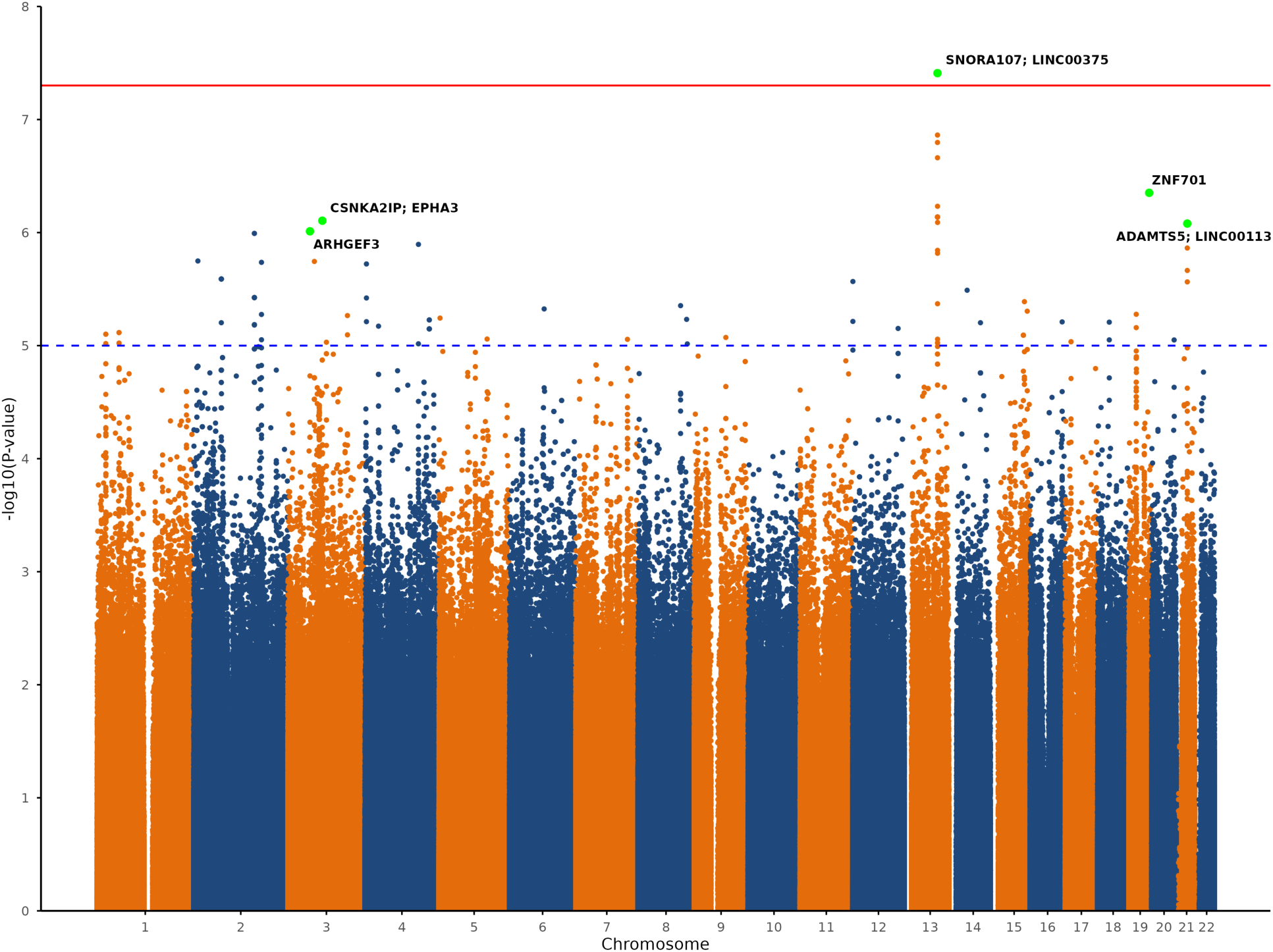
Manhattan plot of genome-wide association results for ACS recurrence in SCD. Each point represents a SNP, ordered by chromosomal position (x-axis) and plotted against the -log₁₀ of its association p-value (y-axis). The red solid horizontal line denotes the genome-wide significance threshold (p = 5 x 10^-8^) and the blue dashed line is the suggestive threshold (p = 1 x 10^-5^). Top 5 independent SNPs have been labelled with their closest genes.

**Table 3:**
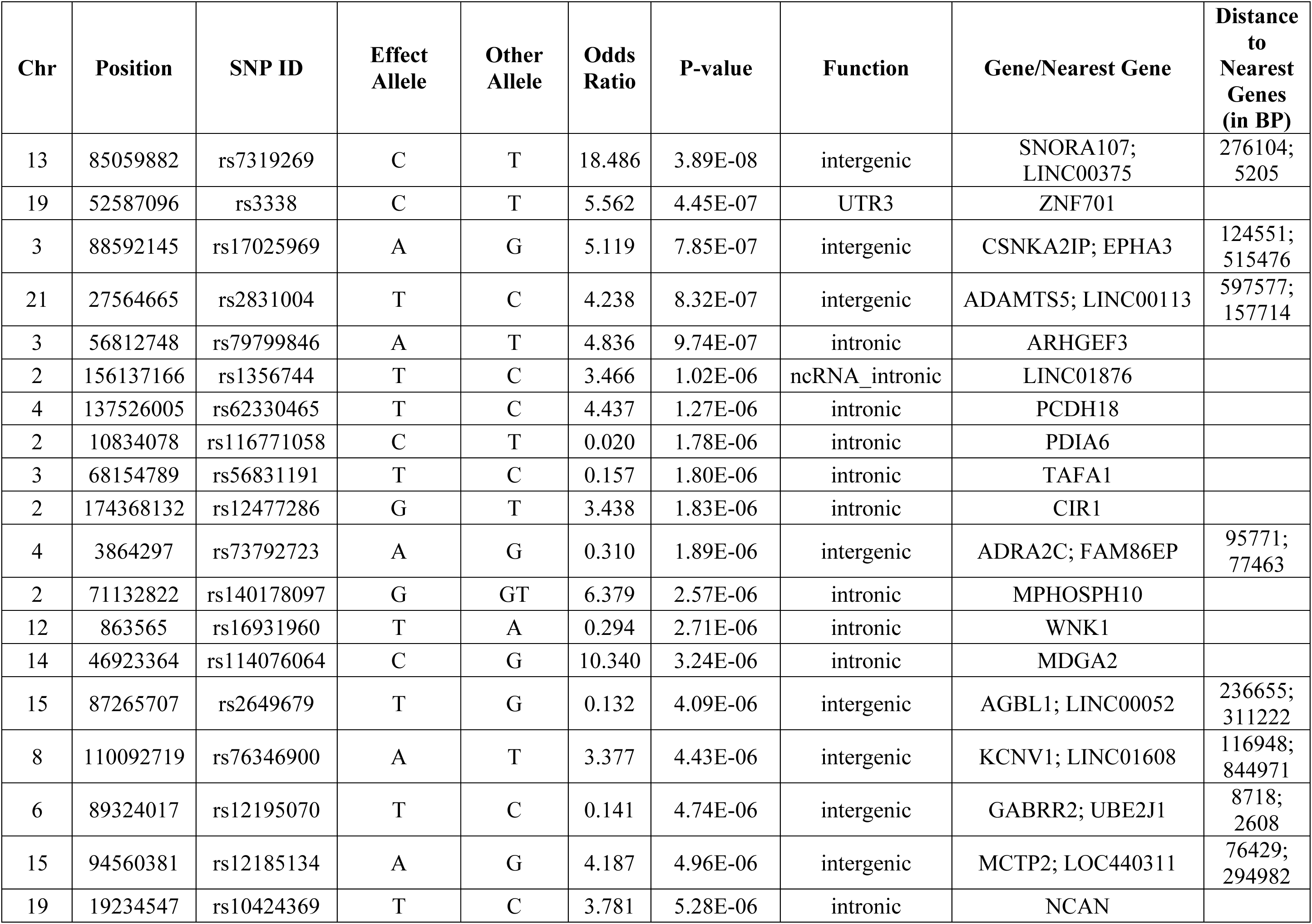
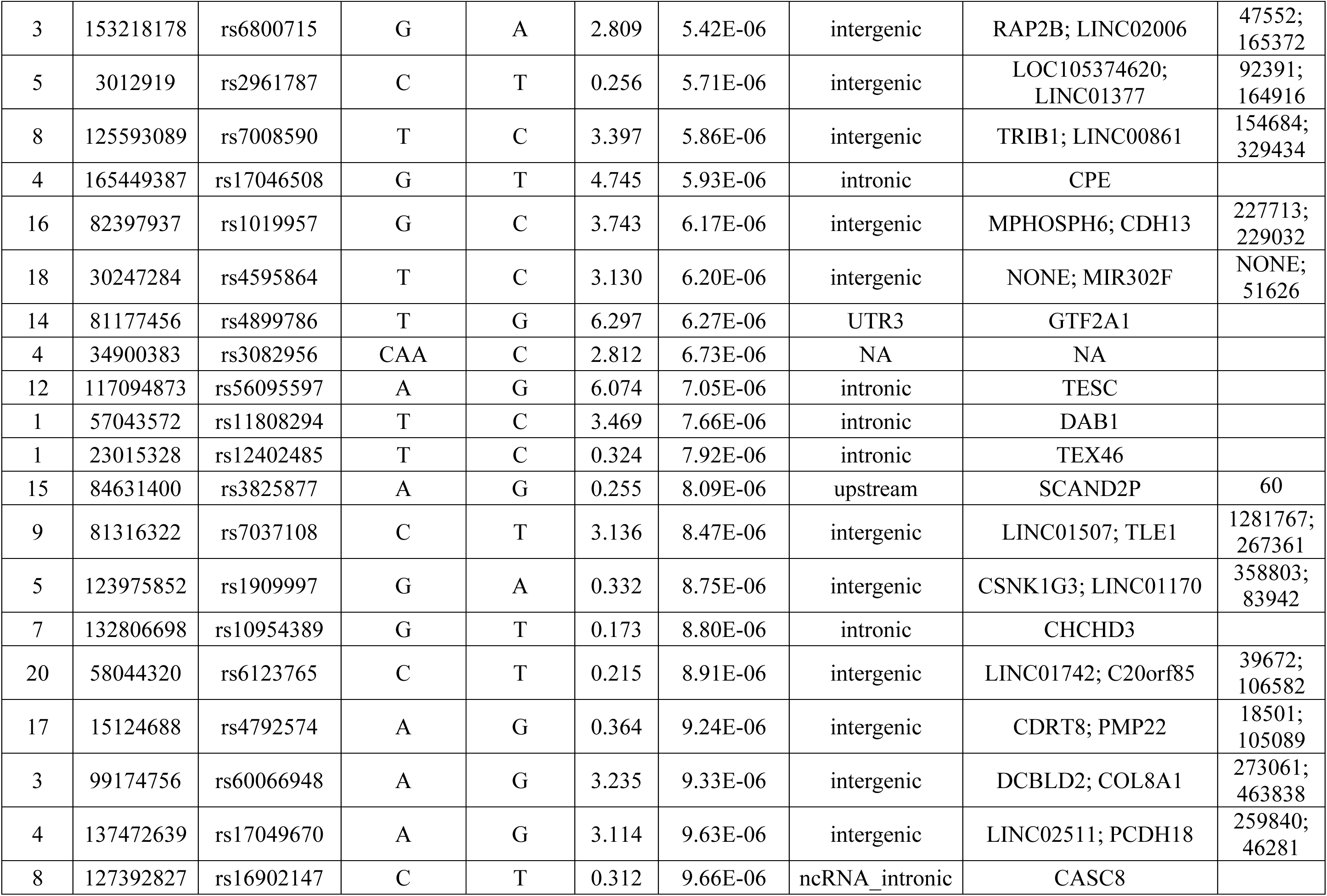
Top SNPs from ACS recurrence GWAS (p < 1 x 10^-5^).

### SNPs associated with recurrent ACS are also associated with protein and metabolite abundance

To understand the functional implications of the top SNPs associated with recurrent ACS episodes, we performed a molecular quantitative trait loci (QTL) analysis. We tested the nominally significant independent SNPs (n=39) against protein (n=1190) and metabolite (n=250) abundance in 185 individuals and 232 individuals, respectively. Age, sex, HU status, and two genotype PCs were included as covariates.

Although no SNP-molecule pair survived FDR correction, 42 SNP-protein pairs had p < 0.001 (Table S2), the strongest being the rs17025969-NCAM1 pair (p = 1.11 x 10^-5^), which we consider hypothesis-generating, given the limited power at this sample size. rs17025969 is an intergenic SNP between *CSNKA2IP* and *EPHA3*, while NCAM1 (neural cell adhesion molecule) is a glycoprotein involved in cell-cell adhesion. Altered expression and activation of cell adhesion molecules is a known primary driver of vaso-occlusive crisis in SCD.^61,62^ EPHA3 is a receptor tyrosine kinase involved in cell-surface signaling and motility. In physiological models, NCAM1 physically and functionally binds to EPHA3, causing autophosphorylation and cytoskeletal remodeling.^63^ Because EPHA3 drives inflammatory cell migration and motility, its signaling axis, alongside NCAM1, could be involved in the systemic inflammation and endothelial adhesion that characterizes ACS and SCD vasculopathy.

Furthermore, to evaluate whether these SNP – protein pairs converge on shared biological processes, we constructed a protein-protein interaction (PPI) network (Figure 4A) comprising 30 nodes and 29 edges, compared with 11 edges expected by chance for a random gene set of equal size (PPI enrichment p = 5.59 x 10^-6^), indicating that these candidate genes are more interconnected than expected under a null model. Gene ontology (GO) cellular component enrichment identified seven significantly enriched terms (Figure 4B), dominated by extracellular and vesicular compartments. Extracellular matrix (GO:0031012) had the highest strength score and enrichment signal (strength = 0.86, signal = 0.82), while extracellular region (GO: 0005576) was the most gene-dense and most significant term (37 genes, FDR = 2.49 x 10^-10^), consistent with a secretory/extracellular signature among the candidate genes. Reactome pathway enrichment identified five significant pathways (Figure 4C), focusing on hemostatic and complement-related processes. Hemostasis (HSA-109582) was the most gene-dense and most significantly enriched pathway (13 genes, strength = 0.89, signal = 0.96, FDR = 1.96 x 10^-5^). Other terms include platelet degranulation; platelet activation, signaling and aggregation; complement cascade; and initial triggering of complement. Together, these results suggest that the candidate genes converge on platelet/hemostatic and complement-mediated pathways.

**Figure 4:**
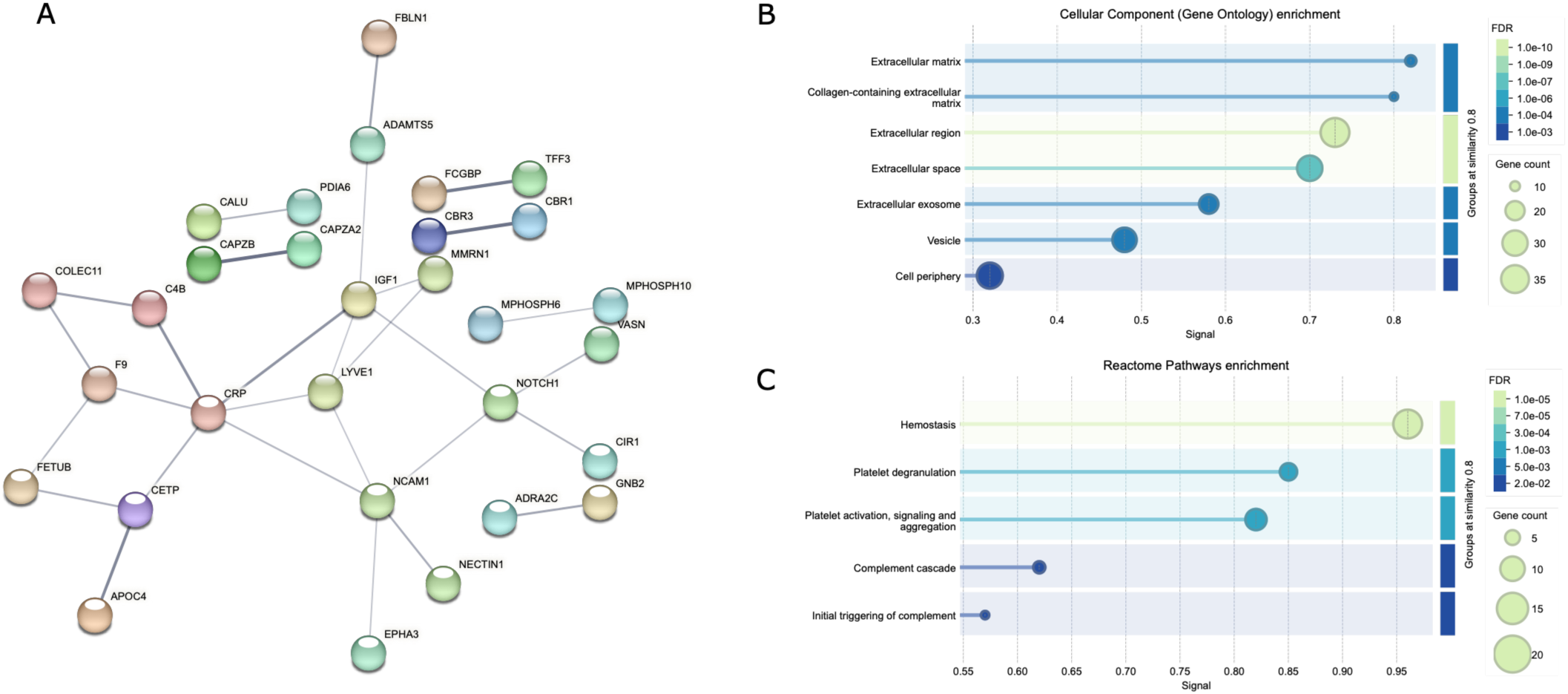
Pathway analysis from SNP-Protein interactions. **(A):** Protein–protein interaction network of candidate genes generated in STRING (evidence view), constructed at a medium confidence threshold (≥ 0.4). Edge thickness denotes the level of confidence based on the amount of supporting evidence (e.g., co-expression, experimentally determined interactions, curated databases, text-mining), per STRING’s standard evidence-channel legend. Genes with no interaction partner at this threshold were hidden for clarity. The network comprises 30 nodes and 29 edges. (**B):** Gene Ontology Cellular Component terms enriched among candidate genes, generated in STRING (terms redundancy-reduced at a similarity threshold of 0.8). Dot position along the x-axis (Strength) reflects log10(observed/expected) gene overlap for each term; dot size reflects the number of candidate genes annotated to that term; dot color reflects statistical significance (FDR-corrected p-value), with lighter tones indicating stronger significance and darker blue indicating weaker (but still significant) enrichment. **(C):** Reactome pathways enriched among candidate genes, generated in STRING. Dot position along the x-axis (Strength) reflects log10(observed/expected) gene overlap for each term; dot size reflects the number of candidate genes annotated to that term; dot color reflects statistical significance (FDR-corrected p-value), with lighter tones indicating stronger significance and darker blue indicating weaker (but still significant) enrichment.

Eight SNP-metabolite pairs were nominally significant (p < 0.001; Table S2), the strongest association being rs12195070 with uridine (p = 1.94 x 10^-4^). rs12195070 is an intergenic SNP between *GABRR2* and *UBE2J1*. Alterations in GABA receptors may play minor regulatory roles in modulating the highly elevated inflammatory pathways and endothelial dysfunction that directly drive ACS.^64^ Notably, the same SNP was also associated with lactic acid, the primary product of anerobic glycolysis. Buildup of lactic acid is a common indication of severe sickling and vaso-occlusive crisis in SCD.^65^

## DISCUSSION

In this study, we identified novel associations of genetic variants with both occurrence and high recurrence of ACS in SCD patients with African ancestry. We further linked recurrence-associated loci to their downstream effects on the plasma proteome and metabolome. By layering network and pathway analysis on top of molecular associations, we identified multiple mechanistic links (including secretory, inflammatory, and hemostatic pathways) between genetic variation and recurrent ACS. This approach helped us infer functional consequences of SNPs, as evident from associations with protein and metabolite abundance, moving beyond associations and providing support for previous work suggesting the roles of coagulation and complement activation in the etiology of ACS.^46,66^

Our survival analysis findings are directionally consistent with the Cooperative Study of Sickle Cell Disease, in which adults with higher ACS episodes experienced higher all-cause mortality^67^, and with the identification of ACS as an independent predictor of early death in the same cohort.^68^ These results also indicate that the association is confined to the highest-burden tail of the ACS distribution rather than to any history of ACS, a distinction consistent with the null association between ACS history (modeled dichotomously) and mortality reported in the UNC adult cohort and with the absence of ACS among the risk factors retained in a recent meta-analysis of North American and European cohorts.^69^ However, given the 53 participants in the highest-burden stratum and the non-significant overall log-rank test, this analysis should be interpreted as hypothesis-generating.

Among the previously reported loci associated with ACS, we were able to marginally replicate one of those — rs2070744 (in *NOS3*)^2,20^ with a p-value of 0.061 — in our recurrence analysis. Thus, most findings presented here are novel ACS associations. The recurrence GWAS identified a largely distinct set of loci from the occurrence analysis, consistent with the clinical observation that recurrence risk is not simply a function of susceptibility to a first event but may also be altered by the consequences of the initial event and intercurrent disease.

The molecular QTL analysis meaningfully extends the interpretation of GWAS results. Among the 42 SNP-protein identified at p < 0.001, associations with CRP, PDE5A, F9, and CO4B, are notable given their established roles in SCD pathophysiology. CRP is a major acute-phase inflammatory biomarker, and elevated levels during or following a crisis are strongly associated with impaired lung function and the clinical severity of ACS.^70–73^ PDE5A plays an important role in the pulmonary vasculature.^74^ Hemolysis lowers circulating plasma NO levels.^75^ This deficiency alters the way that PDE5A processes cellular signals, contributing to complications like pulmonary hypertension, which is a major risk factor and complication with ACS.^75^ Recent proteomic analyses of lung fluids during ACS episodes show localized hyper-activation of the coagulation cascade and the complement system (involving F9 and CO4B).^66^ Based on the results from our functional enrichment analysis and recent evidence, at least part of the genetic risk for ACS recurrence acts through pro-thromboinflammatory pathways and the complement activation system. The SNP-metabolite associations, while fewer in number, point toward the role of lactate and acylcarnitines in SCD. Elevated blood lactate (as a result of anaerobic metabolism) serves as a clinical marker for the severity of tissue hypoxia, acute hemolytic crisis, and impending respiratory complications like ACS.^76^ Chronic hemolysis and endothelial injury induce profound systemic mitochondrial stress and alter fatty acid β-oxidation.^77^ Elevated levels of acylcarnitines correlate with severe SCD phenotypes and the metabolic strain experienced by organs during a severe crisis.^78,79^

This study has important strengths. It highlights the significance of a well-defined ACS phenotype (based on the number of episodes) with distinct cases and controls for identification of risk loci. The use of a two-cohort meta-analysis for the occurrence phenotype improves confidence in our findings beyond a single-cohort analysis. To our knowledge, it is among the first to integrate GWAS, proteomic, and metabolomic data specifically to address ACS recurrence in a SCD cohort.

There are also several limitations of the present study that need to be considered while interpreting our findings. First, the recurrence GWAS and molecular QTL analyses were conducted in a single cohort with a relatively small sample size. This may have hampered our statistical power to detect variants and associations with varying effect sizes, while the lack of phenotypic information on recurrence in Walk-PHaSST prevented replication of the associations. Second, the SNP-protein/metabolite associations reported are suggestive but did not survive correction for multiple testing. Therefore, independent replication for both the recurrence GWAS and the pQTLs and metQTLs is needed. Consistency in the definition of ACS would facilitate genetic or other omic association studies, making comparison across studies more reliable, with confidence that studies have examined the same phenotype.

Future work should also prioritize replication of the identified loci associated with occurrence and recurrence of ACS in independent SCD cohorts with pediatric patients, where the frequency of ACS is higher. Functional validation of the identified variants was beyond the scope of this study; thus, the risk conferred by these variants (through different biological pathways) needs experimental validation. Ultimately, functional validation of the top candidate genes and variants will help to establish a causal link and provide evidence for therapeutic targeting.

Nonetheless, this study provides evidence that ACS recurrence in SCD can be traced to specific genes, proteins, and pathways once genomic and molecular data are integrated. Further, the more extreme phenotype of ACS recurrence may ultimately be a more informative phenotype for analysis and risk prediction. Complications like ACS, which have historically been major contributors to mortality but have eluded from risk prediction, may finally become tractable.

## DATA AVAILABILITY STATEMENT

The whole genome sequence datasets used in this analysis are available via controlled access in the database of Genotypes and Phenotypes (dbGaP) under accession number phs001608 for OMG-SCD and phs001514 for Walk-PHaSST. For questions regarding the OMG-SCD cohort, please contact; for Walk-PHaSST contact.

## ETHICS APPROVAL STATEMENT

Research protocols involving data from the OMG-SCD and Walk-PHaSST patient cohorts were approved by the respective institutional review boards (IRBs) and local ethics committees at all participating enrollment centers.

## PATIENT CONSENT STATEMENT

All participants provided written informed consent prior to enrollment for the use of their clinical, genetic, proteomic, and metabolomic data for future research research purposes.

## CLINICAL TRIAL REGISTRATION

Recruitment of the Walk-PHaSST cohort took place within a clinical trial (NCT00492531) supported by federal funds from the NHLBI, NIH, and Department of Health and Human Services.

## ACKNOWLEDGEMENTS

The authors thank all the subjects for participating in this study. Generation of molecular data for the TOPMed program was supported by NHLBI; whole genome sequencing for OMG-SCD and Walk-PHaSST was performed at Baylor College of Medicine Human Genome Sequencing Center. The TOPMed Informatics Research Centre provided core support including centralized genomic read mapping and genotype calling, along with variant quality metrics and filtering. Core support covering phenotype harmonization, data management, sample-identity QC, and general program coordination was provided by the TOPMed Data Coordinating Centre.

## AUTHORSHIP CONTRIBUTIONS

The research study was designed by AAK and MJT. Walk-PHaSST data were generated by VRG and MTG. Analysis of the data was carried out by AK, MEG, AAK, and MJT. The manuscript was drafted by AK. All authors provided edits to and approved the final version of the manuscript.

