## Supplementary Material for "Multi-omic and Survival Analysis of Acute Chest Syndrome Burden in Sickle Cell Disease"

**Table S1: Top ACS recurrence associated SNP lookups from ACS meta-analysis results.**

| Chr | Position | SNP ID | Effect Allele | Other Allele | Odds Ratio | P-value | Direction in each cohort (OMG, WP) |
|---|---|---|---|---|---|---|---|
| 13 | 85059882 | rs7319269 | C | T | 1.919 | 0.054 | +- |
| 19 | 52587096 | rs3338 | C | T | 1.784 | 0.002 | ++ |
| 3 | 88592145 | rs17025969 | A | G | 2.166 | 1.26E-04 | ++ |
| 21 | 27564665 | rs2831004 | T | C | 1.516 | 0.002 | ++ |
| 3 | 56812748 | rs79799846 | A | T | 1.320 | 0.159 | +- |
| 2 | 156137166 | rs1356744 | T | C | 1.517 | 0.006 | ++ |
| 4 | 137526005 | rs62330465 | T | C | 1.130 | 0.514 | +- |
| 2 | 10834078 | rs116771058 | C | T | 0.611 | 0.027 | -- |
| 3 | 68154789 | rs56831191 | T | C | 0.712 | 0.062 | -+ |
| 2 | 174368132 | rs12477286 | G | T | 1.383 | 0.030 | +- |
| 4 | 3864297 | rs73792723 | A | G | 0.645 | 4.79E-04 | -+ |
| 2 | 71132822 | rs140178097 | G | GT | 1.595 | 0.053 | ++ |
| 12 | 863565 | rs16931960 | T | A | 0.565 | 1.79E-05 | -- |
| 14 | 46923364 | rs114076064 | C | G | 1.644 | 0.071 | +- |
| 15 | 87265707 | rs2649679 | T | G | 0.747 | 0.263 | -+ |
| 8 | 110092719 | rs76346900 | A | T | 1.476 | 0.010 | ++ |
| 6 | 89324017 | rs12195070 | T | C | 0.629 | 0.007 | -- |
| 15 | 94560381 | rs12185134 | A | G | 1.564 | 0.012 | ++ |
| 19 | 19234547 | rs10424369 | T | C | 1.416 | 0.042 | +- |
| 3 | 153218178 | rs6800715 | G | A | 1.103 | 0.447 | +- |
| 5 | 3012919 | rs2961787 | C | T | 0.609 | 0.003 | -+ |
| 8 | 125593089 | rs7008590 | T | C | 1.590 | 0.004 | ++ |
| 4 | 165449387 | rs17046508 | G | T | 2.186 | 9.81E-05 | ++ |
| 16 | 82397937 | rs1019957 | G | C | 1.627 | 0.009 | +- |

| | | | | | | | |
|---|---|---|---|---|---|---|---|
| 18 | 30247284 | rs4595864 | T | C | 1.350 | 0.039 | +- |
| 14 | 81177456 | rs4899786 | T | G | 1.718 | 0.017 | ++ |
| 4 | 34900383 | rs3082956 | CAA | C | 1.317 | 0.030 | +- |
| 12 | 117094873 | rs56095597 | A | G | 1.516 | 0.080 | +- |
| 1 | 57043572 | rs11808294 | T | C | 1.484 | 0.020 | ++ |
| 1 | 23015328 | rs12402485 | T | C | 0.896 | 0.448 | -+ |
| 15 | 84631400 | rs3825877 | A | G | 0.740 | 0.116 | -+ |
| 9 | 81316322 | rs7037108 | C | T | 1.499 | 0.003 | +- |
| 5 | 123975852 | rs1909997 | G | A | 0.633 | 2.17E-04 | -- |
| 7 | 132806698 | rs10954389 | G | T | 0.717 | 0.049 | -+ |
| 20 | 58044320 | rs6123765 | C | T | 0.851 | 0.255 | -+ |
| 17 | 15124688 | rs4792574 | A | G | 0.691 | 0.002 | -- |
| 3 | 99174756 | rs60066948 | A | G | 1.315 | 0.086 | ++ |
| 4 | 137472639 | rs17049670 | A | G | 1.266 | 0.107 | +- |
| 8 | 127392827 | rs16902147 | C | T | 0.688 | 0.004 | -+ |

OMG = OMG-SCD cohort, WP = Walk-PHaSST cohort

**Table S2: Top SNP-molecule pairs associated with top SNPs from ACS recurrence GWAS ($p < 1 \times 10^{-3}$).**

| SNP-Protein Pairs | | | | | |
|---|---|---|---|---|---|
| **SNP ID** | **Protein** | **N** | **Effect** | **P-value** | **FDR** |
| rs17025969 | NCAM1_HUMAN | 185 | 0.310 | 1.11E-05 | 0.515 |
| rs62330465 | KV401_HUMAN | 185 | -0.378 | 2.39E-05 | 0.554 |
| rs3082956 | LV548_HUMAN | 184 | -0.920 | 1.02E-04 | 0.900 |
| rs11808294 | LV140_HUMAN | 185 | -0.306 | 1.17E-04 | 0.900 |
| rs3338 | NECT1_HUMAN | 184 | -0.653 | 1.49E-04 | 0.900 |
| rs17025969 | ADEC1_HUMAN | 185 | 0.760 | 1.59E-04 | 0.900 |
| rs4792574 | THTR_HUMAN | 183 | 0.808 | 1.72E-04 | 0.900 |
| rs2831004 | CETP_HUMAN | 185 | 0.318 | 1.74E-04 | 0.900 |
| rs4899786 | CBR3_HUMAN; CBR1_HUMAN | 114 | -2.021 | 1.82E-04 | 0.900 |
| rs17025969 | APOC4_HUMAN | 185 | -0.480 | 2.89E-04 | 0.900 |
| rs140178097 | CRP_HUMAN | 185 | 1.791 | 3.05E-04 | 0.900 |
| rs4792574 | CAPZB_HUMAN | 183 | 0.684 | 3.13E-04 | 0.900 |
| rs3825877 | IGF1_HUMAN | 185 | -0.552 | 3.24E-04 | 0.900 |
| rs17046508 | TFF3_HUMAN | 185 | -0.591 | 3.65E-04 | 0.900 |
| rs4792574 | MMRN1_HUMAN | 185 | 0.467 | 3.91E-04 | 0.900 |
| rs3338 | SBSN_HUMAN | 180 | -0.971 | 3.92E-04 | 0.900 |
| rs116771058 | FCGBP_HUMAN | 185 | 0.918 | 3.93E-04 | 0.900 |
| rs60066948 | FETUB_HUMAN | 185 | -0.294 | 4.43E-04 | 0.900 |
| rs12477286 | HINT1_HUMAN | 184 | -0.655 | 5.17E-04 | 0.900 |
| rs62330465 | HV439_HUMAN | 185 | -0.279 | 5.19E-04 | 0.900 |
| rs17025969 | NOTC1_HUMAN | 181 | 0.826 | 5.90E-04 | 0.900 |
| rs4899786 | LYVE1_HUMAN | 185 | -0.481 | 6.00E-04 | 0.900 |
| rs7319269 | FN3K_HUMAN | 185 | -1.497 | 6.20E-04 | 0.900 |
| rs17025969 | FBLN1_HUMAN | 185 | 0.336 | 7.32E-04 | 0.900 |
| rs17025969 | HV320_HUMAN | 185 | 0.370 | 7.38E-04 | 0.900 |
| rs60066948 | CALU_HUMAN | 175 | -0.970 | 7.44E-04 | 0.900 |
| rs12477286 | PDE5A_HUMAN | 185 | -0.431 | 7.87E-04 | 0.900 |
| rs12477286 | NB5R1_HUMAN | 168 | -0.981 | 7.88E-04 | 0.900 |
| rs4792574 | BAP31_HUMAN | 134 | 0.980 | 7.91E-04 | 0.900 |

| | | | | | |
|---|---|---|---|---|---|
| rs2649679 | JKAMP_HUMAN | 56 | 5.162 | 7.96E-04 | 0.900 |
| rs1019957 | SIRPG_HUMAN | 185 | 0.832 | 8.09E-04 | 0.900 |
| rs17025969 | VASN_HUMAN | 185 | 0.295 | 8.26E-04 | 0.900 |
| rs73792723 | COL11_HUMAN | 185 | 0.160 | 8.78E-04 | 0.900 |
| rs17049670 | FN3K_HUMAN | 185 | -0.688 | 8.80E-04 | 0.900 |
| rs2649679 | FA9_HUMAN | 185 | -0.272 | 8.92E-04 | 0.900 |
| rs4792574 | ENDD1_HUMAN | 185 | 0.488 | 9.09E-04 | 0.900 |
| rs17025969 | LG3BP_HUMAN | 185 | 0.563 | 9.15E-04 | 0.900 |
| rs4792574 | CAZA2_HUMAN | 185 | 0.428 | 9.16E-04 | 0.900 |
| rs17049670 | HV313_HUMAN | 185 | -0.522 | 9.16E-04 | 0.900 |
| rs17025969 | CO4B_HUMAN | 185 | -0.354 | 9.17E-04 | 0.900 |
| rs11808294 | CBPN_HUMAN | 185 | 0.149 | 9.58E-04 | 0.900 |
| rs4792574 | GBB2_HUMAN | 177 | 0.640 | 9.79E-04 | 0.900 |
| **SNP-Metabolite Pairs** | | | | | |
| **SNP ID** | **Metabolite (HMDB ID)** | **N** | **Effect** | **P-value** | **FDR** |
| rs12195070 | Uridine (HMDB00296) | 232 | 0.431 | 1.94E-04 | 0.658 |
| rs3338 | Decanoylcarnitine (HMDB00651) | 232 | 0.513 | 2.89E-04 | 0.658 |
| rs3338 | Octanoylcarnitine (HMDB00791) | 232 | 0.561 | 2.90E-04 | 0.658 |
| rs12195070 | Lactic acid (HMDB00190) | 232 | 0.485 | 4.16E-04 | 0.658 |
| rs12195070 | Choline (HMDB00097) | 232 | 0.490 | 6.56E-04 | 0.658 |
| rs3338 | Hexanoylcarnitine (HMDB0000756) | 232 | 0.394 | 6.58E-04 | 0.658 |
| rs3338 | trans-2-Dodecenoylcarnitine (HMDB13326) | 232 | 0.428 | 6.60E-04 | 0.658 |
| rs16931960 | D-Glucose (HMDB00122) | 232 | -0.603 | 7.57E-04 | 0.660 |

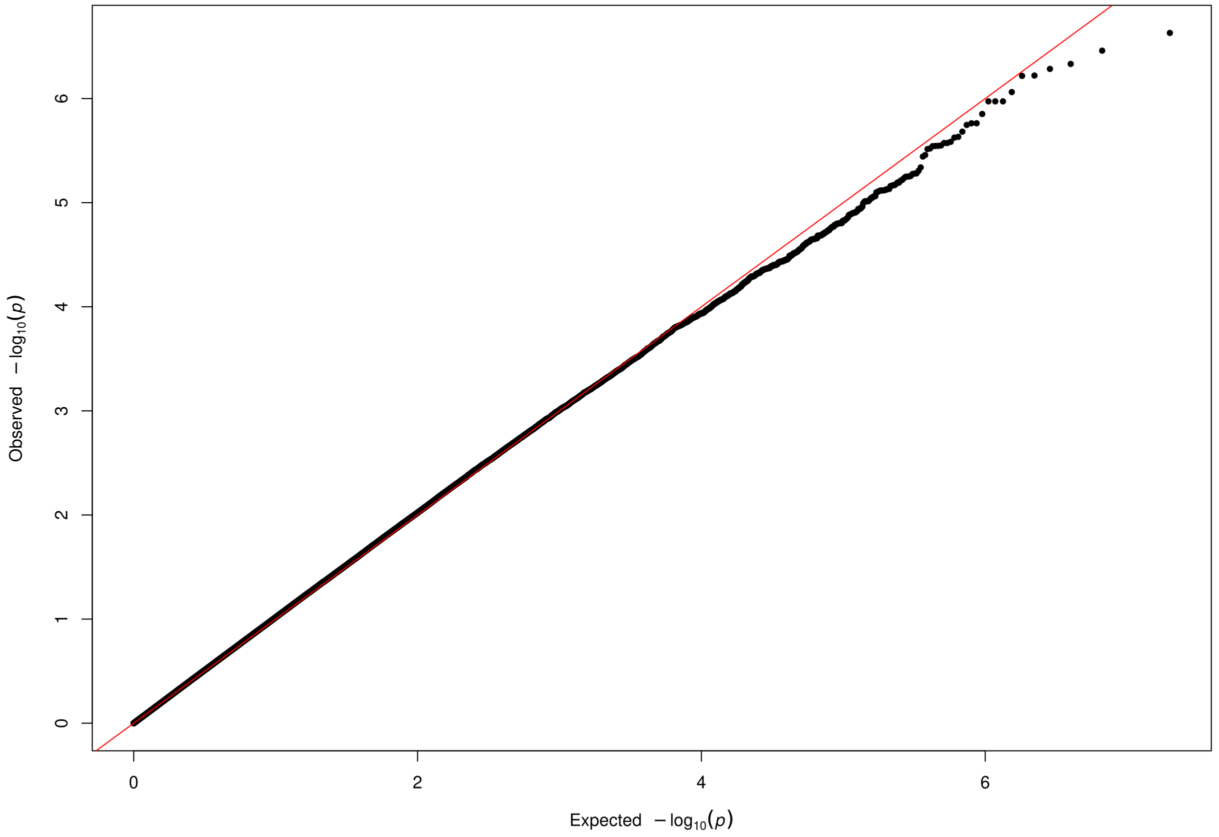


**Figure S1:** Quantile–quantile plot of meta-analysis association results for ACS occurrence in two SCD cohorts. Observed $-\log_{10}$ meta-analysis p-values (y-axis) are plotted against the null expectation (x-axis), with the red diagonal indicating no association. The meta-analysis genomic inflation factor was $\lambda = 1.03$.

A

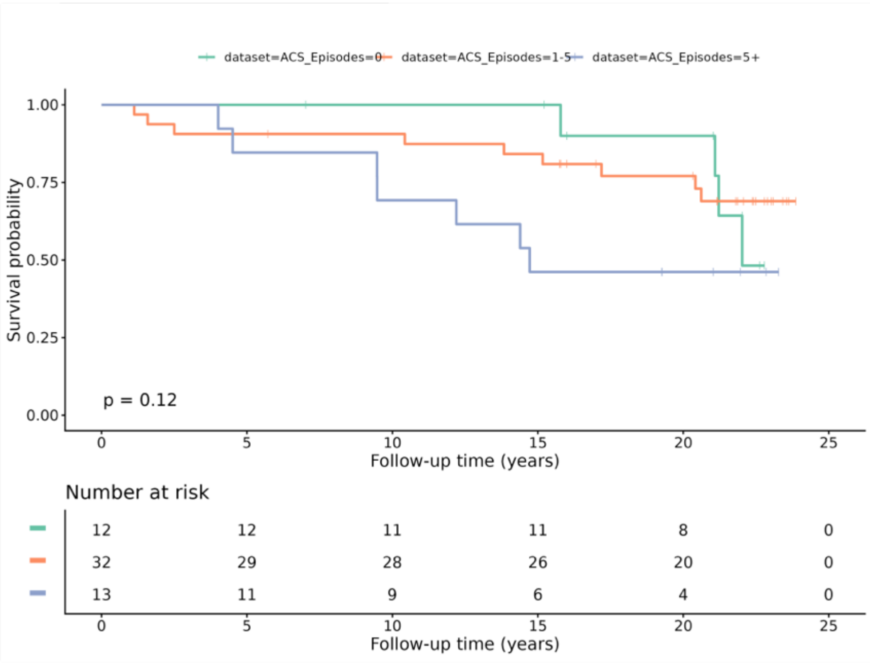

dataset=ACS_Episodes=0
dataset=ACS_Episodes=1-5
dataset=ACS_Episodes=5+
Survival probability
1.00
0.75
0.50
0.25
0.00
p = 0.12
0 5 10 15 20 25
Follow-up time (years)
Number at risk
12 12 11 11 8 0
32 29 28 26 20 0
13 11 9 6 4 0
0 5 10 15 20 25
Follow-up time (years)


B

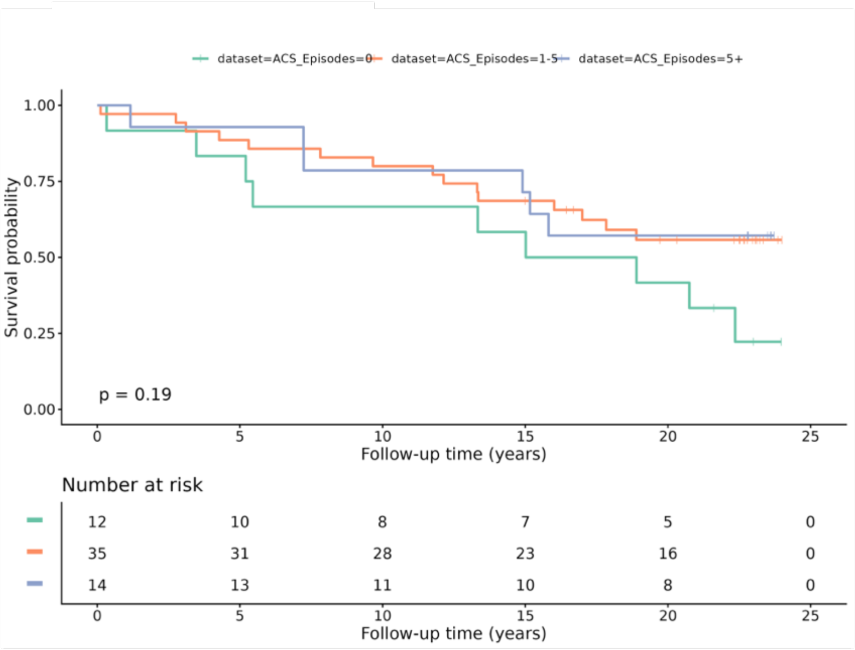

dataset=ACS_Episodes=0
dataset=ACS_Episodes=1-5
dataset=ACS_Episodes=5+
Survival probability
1.00
0.75
0.50
0.25
0.00
p = 0.19
0 5 10 15 20 25
Follow-up time (years)
Number at risk
12 10 8 7 5 0
35 31 28 23 16 0
14 13 11 10 8 0
0 5 10 15 20 25
Follow-up time (years)


C

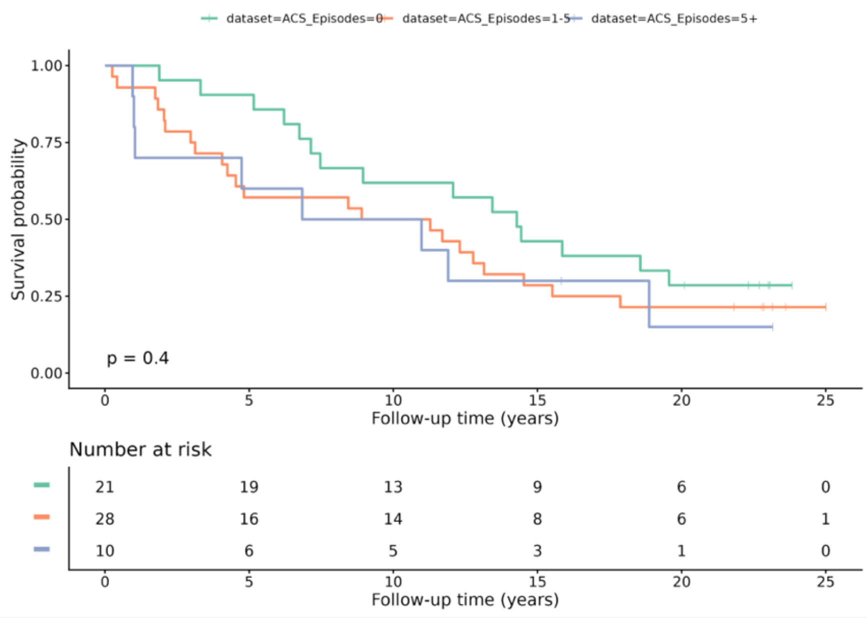

dataset=ACS_Episodes=0
dataset=ACS_Episodes=1-5
dataset=ACS_Episodes=5+
Survival probability
1.00
0.75
0.50
0.25
0.00
p = 0.4
0 5 10 15 20 25
Follow-up time (years)
Number at risk
21 19 13 9 6 0
28 16 14 8 6 1
10 6 5 3 1 0
0 5 10 15 20 25
Follow-up time (years)

**Figure S2**: Survival curves for OMG-SCD participants within quantiles of age at examination (quantile 1 in Figure 2B): **(A)** quantile 2, 23.0-31.0 years (n=12, 32, and 13); **(B)** quantile 3, 31.0-41.0 years (n=12, 35, and 14); **(C)** quantile 4, 41.0-70.0 years (n=21, 28, and 10). In all plots, vertical tick marks denote censored observations and the table beneath each plot gives the number remaining at risk at 5-year intervals. Log-rank p-values are also shown within each plot and were not adjusted for multiple comparisons.

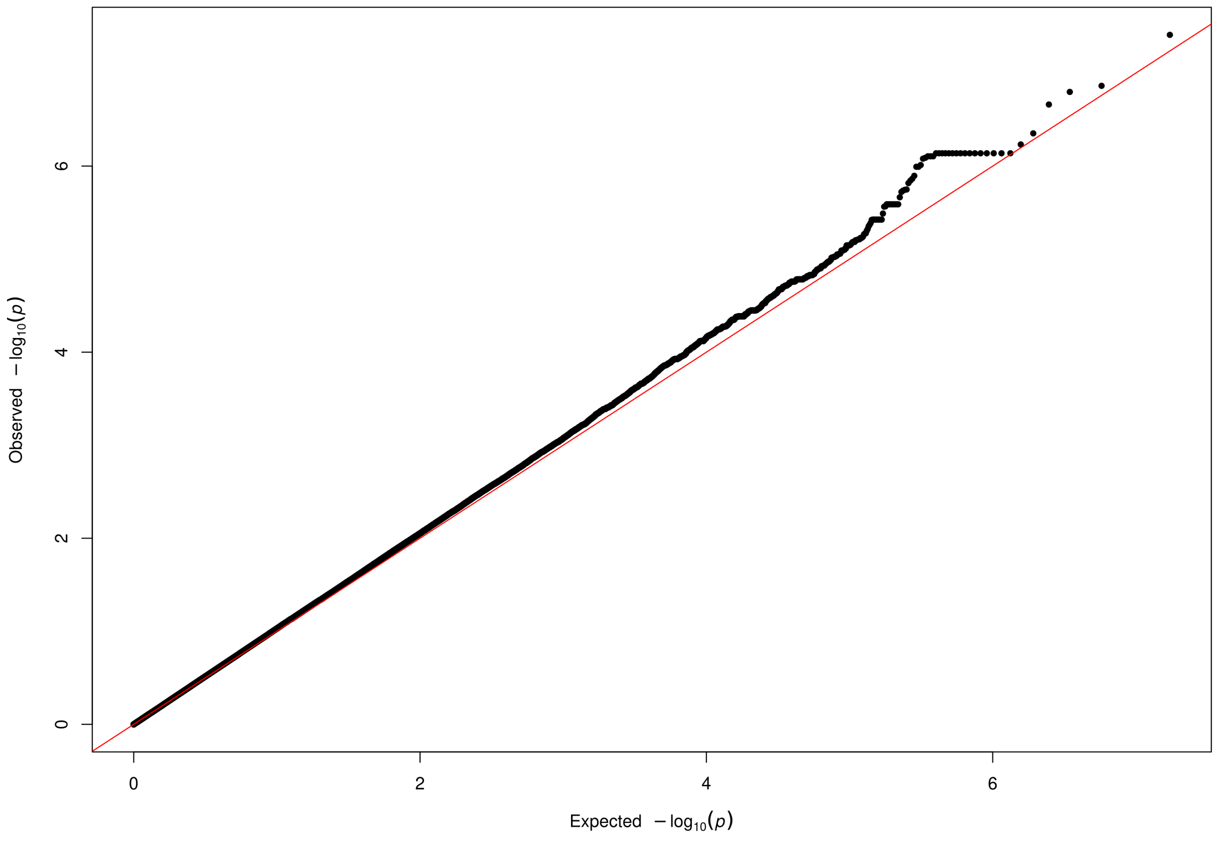


**Figure S3:** Quantile–quantile plot of genome-wide association results for ACS recurrence in SCD. Observed $-\log_{10}$ p-values (y-axis) are plotted against those expected under the null hypothesis of no association (x-axis). The red diagonal line represents the null expectation. The genomic inflation factor was $\lambda = 1.04$.
